# Different contribution of missense and loss-of-function variants to the genetic structure of familial and sporadic Meniere disease

**DOI:** 10.1101/2025.04.22.25326157

**Authors:** Alberto M. Parra-Perez, Alvaro Gallego-Martinez, Alba Escalera-Balsera, Paula Robles-Bolivar, Patricia Perez-Carpena, Jose A. Lopez-Escamez

## Abstract

Meniere disease (MD) is a chronic inner ear disorder with significant heritability. This study aims to compare the burden of rare high-and moderate-impact protein-coding variants in a MD cohort to determine whether the genetic burden in sporadic MD (SMD) overlaps familial MD (FMD), potentially revealing hidden inheritance in SMD.

In this study exome sequencing identified rare variants in unrelated FMD (N=93) and SMD (N=287) patients. Gene Burden Analysis (GBA) was performed, and candidate genes were prioritized using the number of individuals with variants, inner-ear gene expression, and hearing and balance-related phenotypic annotations.

In FMD patients, a higher accumulation of missense and loss-of-function variants was observed compared to SMD, particularly in genes associated with auditory and vestibular functions. GBA identified 269 enriched genes in SMD, with 31 annotated for inner ear phenotypes, while FMD had 432 genes with 51 pinpointed. Sporadic and FMD overlapped in 28.1% of the enriched genes, with *ADGRV1*, *MEGF8* and *MYO7A* the most commonly found.

In conclusion, SMD and FMD have a divergent genetic architecture. Both SMD and FMD have an overload of missense variants in stria vascularis and hair cell stereocilia genes that suggests different mechanisms in MD pathogenesis and a multiallelic-recessive inheritance pattern.

## Introduction

Meniere Disease (MD) is a chronic, progressive inner ear disorder characterized by episodic vertigo associated with a fluctuating sensorineural hearing loss (SNHL), tinnitus or aural fullness^1^. MD prevalence is population-specific, being more prevalent in Europeans than in Asians ^2^. The MD phenotype is variable and several clinical subgroups have been defined according to its familial aggregation and associated comorbidities, such as migraine, and autoimmune diseases ^3^. In addition, an important number of MD patients show a systemic proinflammatory response, which is associated either with high levels of IgE and type 2 immune response and/or high levels IL1ß ^4^.

It is considered a complex disorder with a strong familial aggregation in Europeans ^5^. Familial MD (FMD) was initially reported as an autosomal dominant condition with incomplete penetrance and anticipation in subsequent generations ^6^, but patients could also show a compound recessive ^7^ or digenic inheritance pattern ^8^. Exome sequencing (WES) has identified several genes related to FMD. Single nucleotide variants (SNV) in *DTNA* (HGNC:3057), *FAM136A* (HGNC:25911), *PRKCB* (HGNC:9395), *DPT* (HGNC:3011), *SEMA3D* (HGNC:10726), *COCH* (HGNC:2180), *GUSB* (HGNC:4696), and *SLC6A7* (HGNC:11054) were identified in different families with an autosomal dominant inheritance pattern with incomplete penetrance. Likewise, genes with a recessive inheritance pattern such as *HMX2* (HGNC:5018), *LSAMP* (HGNC:6705) and *STRC* (HGNC:16035) have been reported. Nevertheless, no replication of these findings has been found either in other FMD or Sporadic MD (SMD) cases, suggesting a considerable genetic heterogeneity ^9^.

Since these genes were being discovered in individual families, the Gene Burden Analysis (GBA) has allowed the identification of rare variants in the same genes in a broader number of families. This approach has led to the discovery of a burden of rare variation in three SNHL genes in several MD families, which are *OTOG* (HGNC:8516), *MYO7A* (HGNC:7606), and *TECTA* (HGNC:11720) ^7,8,10^, with a recessive, digenic and dominant inheritance pattern, respectively.

Although significant progress has been made in identifying variants and genes associated with FMD, the genetic underpinnings of SMD remain less studied. Despite the absence of an apparent hereditary pattern in SMD, previous studies reported an enrichment of missense variants in SNHL genes (*GJB2* (HGNC:4284), *USH1G* (HGNC:16356), *SLC26A4* (HGNC:8818), *ESRR* (HGNC:3473), and *CLDN14* (HGNC:2035)) ^11^ and genes related to axonal guidance signaling (*NTN4* (HGNC:13658)) ^12^.

In this study, we aimed to compare the distribution of rare variants with a high and moderate impact on the protein in a large cohort of MD patients. In addition, we sought to ascertain whether the genetic burden of SMD diverges or overlaps with FMD, to uncover hidden inheritance in sporadic cases.

## Results

### Genetic variants in Sporadic and Familiar MD

The total number of genetic variants obtained was 195,454 and 330,037 in the FMD cohort (N=93) and SMD cohort (N=287 patients) (Table S1), respectively. The distribution of variants with an AF < 0.01 was similar between FMD and SMD (Table S2).

Based on the number of missense variants with an AF < 0.01 per patient, FMD patients showed a significantly higher variant accumulation in all genes (FMD median (IQR) = 2410 (2369.25, 2445.75); SMD median (IQR) = 2214 (2184.5, 2246); p = 6.52×10^-45^), in genes related to an auditory vestibular phenotype (FMD median (IQR) = 226.5 (219.75, 236); SMD median (IQR) = 194 (184, 201); p = 2.42×10^-43^), genes encoding hair cell stereocilia and tectorial membrane (TM) proteins (FMD median (IQR) = 122 (115, 129); SMD median (IQR) = 103 (96.5, 109); p = 1.472×10^-32^), and genes expressed in the stria vascularis (FMD median (IQR) = 1179 (1156. 75, 1200.50); SMD median (IQR) = 1057 (1037, 1077); p = 1.49×10^-45^), compared to SMD patients (Fig. 1a).

**Fig 1.**
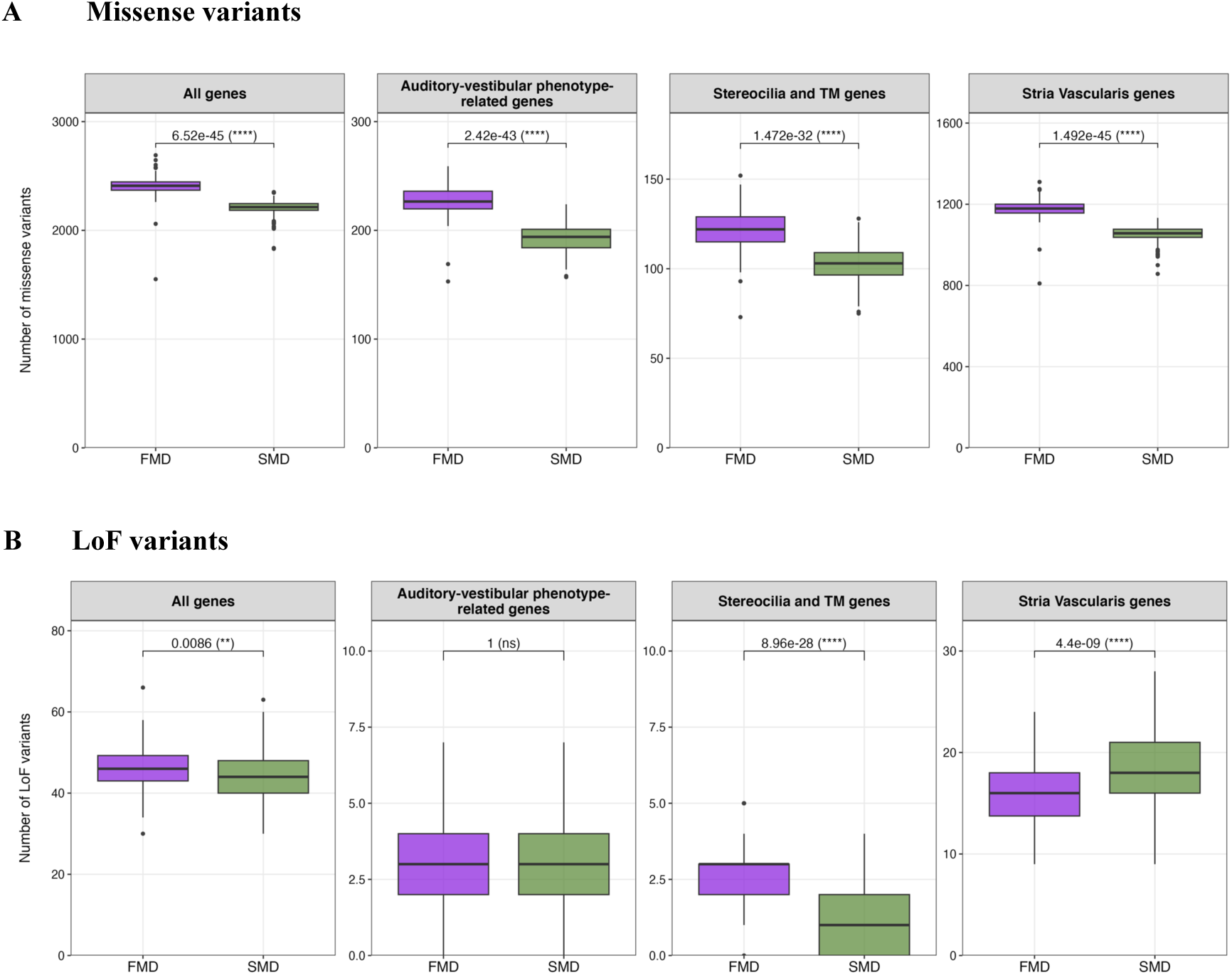
Boxplot comparing the number of missense (**A**) and loss of function (LoF) (**B**), variants in FMD (purple) and SMD (green) patients. The number of total variants in all exome genes, in genes previously associated with an auditory-vestibular phenotype in HPO or MGI, genes expressing proteins present in hair cell stereocilia and tectorial membrane (TM) and genes expressed in Stria vascularis are compared from left to right. The Bonferroni-corrected p-value obtained with the Mann-Whitney test is shown. p-value < 0.05 is considered as significant. NS, not significant.

On the other hand, the number of loss-of-function (LoF) variants with an AF < 0.01 was higher in FMD compared to SMD considering all genes (FMD median (IQR) = 46 (43, 49.25); SMD median (IQR) = 44 (40, 48); p = 8.6×10^-3^) and genes encoding stereocilia and TM proteins (FMD median (IQR) = 3 (2, 3); SMD median (IQR) = 1 (0, 2); p = 8.96×10^-28^). Nevertheless, the accumulation of LoF variants in FMD and SMD is similar for genes associated with an auditory and vestibular phenotype and significantly lower for genes expressed in stria vascularis (FMD median (IQR) = 16 (13.75, 18); SMD median (IQR) = 18 (16, 21); p = 4.4×10^-9^) (Fig. 1b).

### Enriched genes in Sporadic MD

In the SMD cohort, 74,097 variants with an AF < 0.01 and a high or moderate impact on the protein, according to VEP, were found in 15,709 genes. Significant variant enrichment in 779 genes (Table S3), with 8,145 variants, was found in the GBA compared to the reference populations (FDR < 0.05). After filtering FLAGS genes and considering genes with variants present in more than 3 % of the individuals (N > 8 patients), a total of 269 genes (Table S4), with 4,272 variants (Table S5), were obtained. Given the large number of potential candidate genes, those with HPO or MGI phenotypic annotations related to the inner ear were prioritized. This resulted in a total of 31 genes (Table S6), with 538 variants (Table S7), among which 7 genes with 151 variants are included in the OTOscope v9 panel used for genetic hearing loss diagnosis (Table S6) (Fig 2a,b,c,d). Additionally, 11 of these genes are expressed in hair cell stereocilia (Table S6). Most of the variants identified in the 31 genes were missense (496 variants), with 12 affecting splicing regions. Additionally, 9 frameshift, 4 stop-gained, 8 in-frame insertion, and 19 in-frame deletion variants were found. Notably, 9 different patients carried the following variants: NC_000016.10:g.88431891C>T in *ZNF469* (HGNC:26216), and NC_000010.11:g.53961876C>A in *PCDH15* (HGNC:14674).

**Fig 2.**
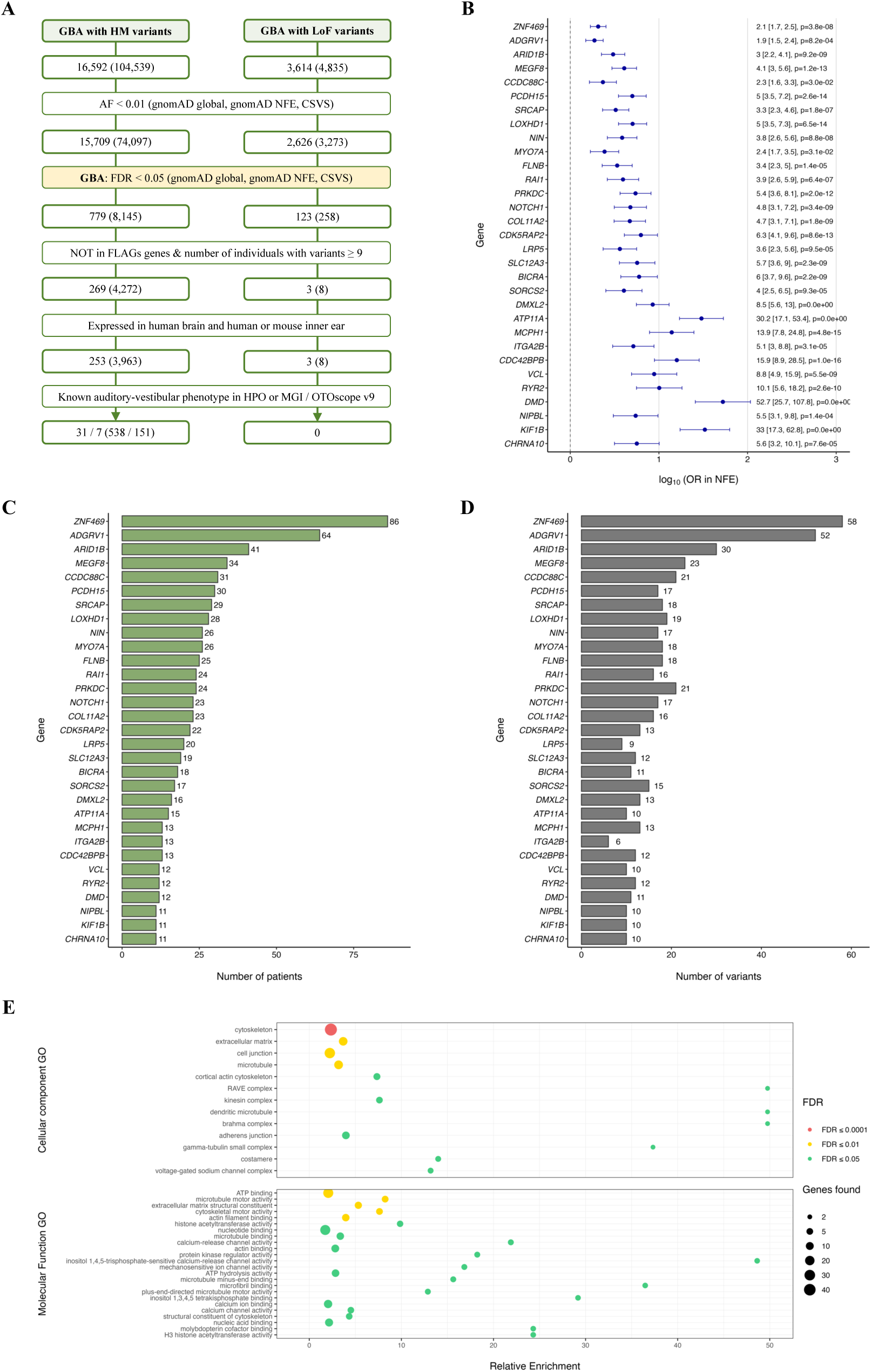
Rare variant analysis and functional enrichment results of candidate genes for sporadic MD cohort. **A**: Flow chart summarizing the prioritization strategy and the result of the GBA for SMD patients, variants filtered by allelic frequency (AF) < 0.01. **B**: Odds ratio (OR) and 95% CI, expressed as log10, of the genes associated with an auditory-vestibular phenotype enriched in variants with a High or Moderate (HM) impact on the protein and LoF, filtered by AF < 0.01, in the SMD cohort, against gnomAD NFE. Genes were ranked according to the number of individuals with variants. **C**: Histogram of the number of SMD patients and **(D)** number of variants found in genes associated with an auditory-vestibular phenotype enriched in HM and LoF variants. **E**: Dot plot representing relative enrichment, FDR-adjusted p-value and number of genes associated with each term using the enriched genes in the SMD cohort expressed in the inner ear. Enrichment analysis was performed using Cellular Components data from the Gene Ontology (GO) database (top) and Molecular Functions data from the GO database (bottom). Red color represents p-value < 0.0001; orange, p-value < 0.001; yellow, p-value < 0.01; and green p-value < 0.05. CSVS: Collaborative Spanish Variant Server, Spanish population; FDR: p-value corrected by False Discovery Ratio; FLAGS: FrequentLy mutAted GeneS; gnomAD NFE: Non-Finnish European for gnomAD; gnomAD: Global population for gnomAD; HPO: Human Phenotype Ontology; MGI: Mouse Genome Informatics

A total of 270 patients (94% of the SMD cohort) had variants in the 31 enriched genes. Among them, 58 individuals had a unique variant, while the majority had at least two variants in the enriched genes, up to a maximum of 8 genes with a variant in one individual. Furthermore, 11 patients had at least 2 variants in *ADGRV1* (HGNC:17416), 5 patients had 2 variants in *MEGF8* (HGNC:3233), 3 in *MYO7A* (HGNC:7606), and 3 patients with 2 variants in *FLNB* (HGNC:3755), *KIF1B* (HGNC:16636), and *PRKDC* (HGNC:9413).

Accounting for possible digenic inheritance, 31 enriched gene pairs with variants were identified in SMD patients. Among these, the *ADGRV1*-*MYO7A* pair, which resultant proteins physically interact in hair cell stereocilia, was found in 7 patients.

Considering only LoF variants, 3,273 LoF variants were found in 2,626 genes with a MAF<0.01. GBA identified 123 genes (Table S8) with 258 variants significantly (FDR<0.05) enriched in the SMD cohort. After applying the aforementioned filters, 3 genes (Table S9), with 8 variants (Table S10), were defined as candidate genes. The enriched genes, ranked by the number of individuals with variants, were: *SYNGAP1* (HGNC:11497), *SKA3* (HGNC:20262), and *BTNL8* (HGNC:26131). None of these genes have phenotypic annotations related to the inner ear in HPO and MGI.

### Enriched genes in Familiar MD

A total of 28,920 variants, with an AF < 0.01 and high or moderate impact, across 11,823 genes were found in the FMD cohort. GBA showed significant enrichment in 740 genes (Table S11) with 4,073 variants. By excluding FLAGs and genes with variants in less than 3 subjects, 432 genes (Table S12) with 2,694 variants were reported (Table S13). The prioritization of genes with inner ear phenotypic annotations yielded 51 genes (Table S14), with 312 variants (Table S15), including 11 genes with 86 variants in the OTOscope v9 panel (Table S14) (Fig 3a,b,c,d). Moreover, 13 of these genes were expressed in hair cell stereocilia (Table S14). Most of the variants found in the 51 enriched genes were missense (282 variants), 9 of which affected splice regions. In addition, 6 frameshift variants, 2 start lost, 1 stop lost, 2 splice acceptor, 1 splice donor, 7 in-frame deletion and 1 in-frame insertion were found. The variants shared by a higher number of patients, 4 for each variant, were NC_000005.10:g.90706355A>C in *ADGRV1* (HGNC:17416), NC_000009.12:g.136505868G>A in *NOTCH1* (HGNC:7881) gene NC_000019.10:g.47698710G>A in *BRICA*(HGNC:4332), NC_00006.10:g.1952959C>T in *RPL3L* (HGNC:10351), NC_000017.11:g.18278232G>A in *TOP3A* (HGNC:11992), and NC_000019.10:g.50266974G>A in *MYH14* (HGNC:23212).

**Fig 3.**
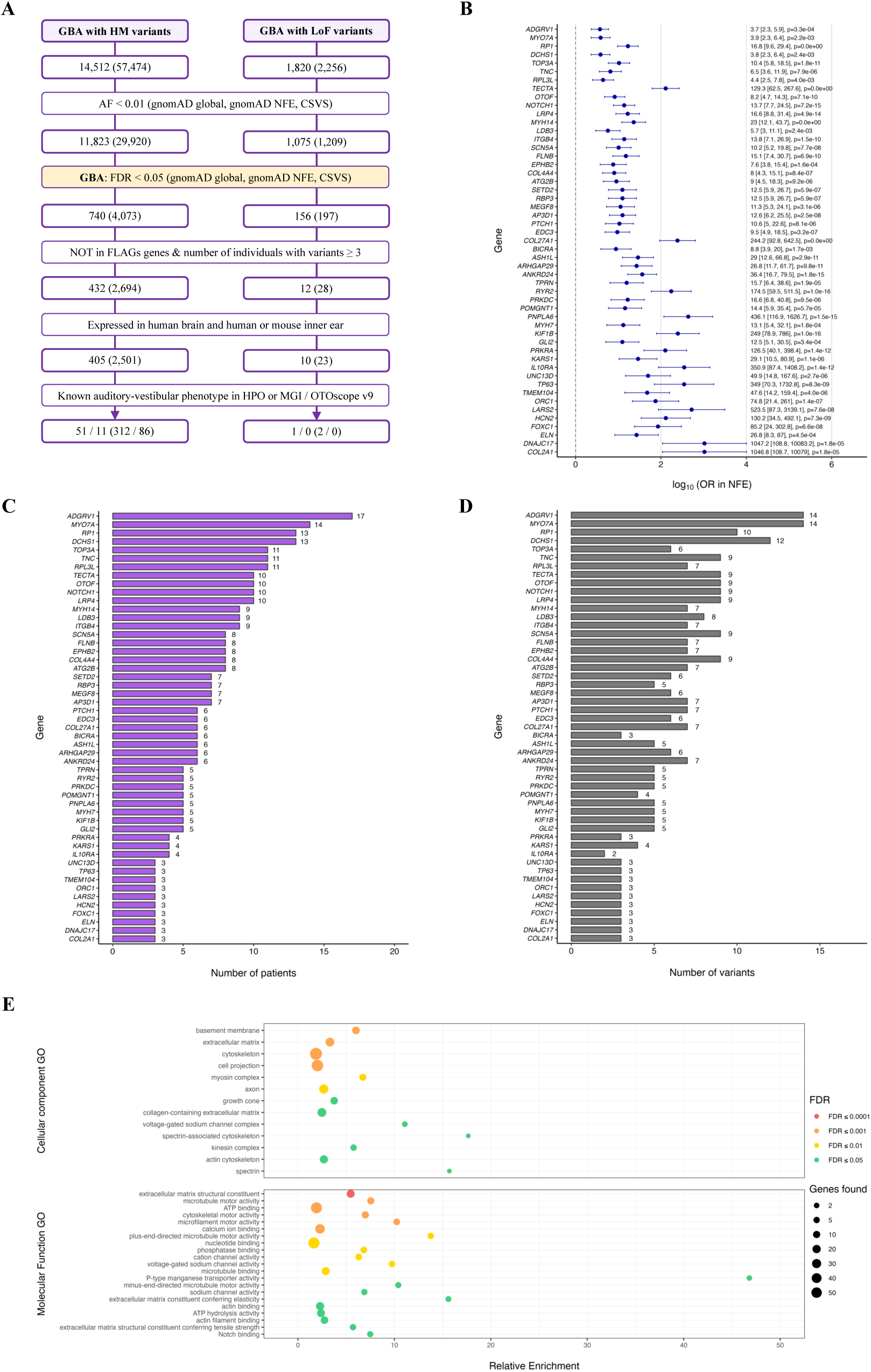
Rare variant analysis and functional enrichment results of candidate genes for familiar MD cohort. **A**: Flow chart summarizing the prioritization strategy and the result of the GBA for FMD patients, variants filtered by allelic frequency (AF) < 0.01. **B**: Odds ratio (OR) and 95% CI, expressed as log10, of the genes associated with an auditory-vestibular phenotype enriched in variants with a High or Moderate (HM) impact on the protein and LoF, filtered by AF < 0.01, in the FMD cohort, against gnomAD NFE. Genes were ranked according to the number of individuals with variants. **C**: Histogram of the number of FMD patients and **(D)** number of variants found in genes associated with an auditory-vestibular phenotype enriched in HM and LoF variants. **E**: Dot plot representing relative enrichment, FDR-adjusted p-value and number of genes associated with each term using the enriched genes in the FMD cohort expressed in the inner ear. Enrichment analysis was performed using Cellular Components data from the Gene Ontology (GO) database (top) and Molecular Functions data from the GO database (bottom). Red color represents p-value < 0.0001, orange p-value < 0.001, yellow p-value < 0.01 and green p-value < 0.05. CSVS: Collaborative Spanish Variant Server, Spanish population; FDR: p-value corrected by False Discovery Ratio; FLAGS: FrequentLy mutAted GeneS; gnomAD NFE: Non-Finnish European for gnomAD; gnomAD: Global population for gnomAD; HPO: Human Phenotype Ontology; MGI: Mouse Genome Informatics

It should be noted that a total of 89 patients (95.7% of the FMD cohort) had variants in the 51 enriched genes. Among these, everyone had two variants in the same or different enriched genes, and 71 individuals had 3 or more variants, with one patient having up to 9 variants.

Finally, regarding LoF variants with a MAF < 0.01, a total of 1,324 variants in 1,075 genes were found in the FMD cohort. After performing GBA, 156 genes (Table S16) with 197 variants were significantly enriched. After filtering steps, including only expressed genes in the inner ear and brain, 10 genes (Table S17) with 23 variants (Table S18) could be considered as potential candidates in the FMD group. The enriched genes, ranked by number of patients with variants, were: *SKA3* (HGNC:20262), *BTNL8* (HGNC:26131), *RBM5* (HGNC:9902), *SYNGAP1* (HGNC:11497), *MYO15B* (HGNC:14083), *LPIN3* (HGNC:14451), *CASP10* (HGNC:1500)*, LRP8* (HGNC:6700), *PRKRA* (HGNC:9438), and *SYTL2* (HGNC:15585). The *PRKRA* gene could be highlighted since in the MGI database it is associated with an increased or absent threshold for auditory brainstem response (MP:0011967) and phenotypes related to ear development and temporal bone morphology.

### Shared enriched genes in Familiar and Sporadic MD

A total of 71 enriched genes were shared in SMD and FMD. In this context, considering the enriched genes, expressed in inner ear and brain, with high and moderate impact variants, 28.1% (71/253) and 17.5% (71/405) of the genes were shared between SMD and FMD (Fig. 4a). Considering genes with phenotypic annotations related to the inner ear, 29% (9/31) and 17.6% (9/51) of the enriched genes were shared for SMD and FMD, respectively (Fig. 4b). The common enriched genes with phenotypic annotation were: *ADGRV1*, *BICRA*, *FLNB* (HGNC:3755), *KIF1B* (HGNC:16636), *MEGF8* (HGNC:3233), *MYO7A*, *NOTCH1*, *PRKDC* (HGNC:9413), and *RYR2* (HGNC:10484).

**Fig 4.**
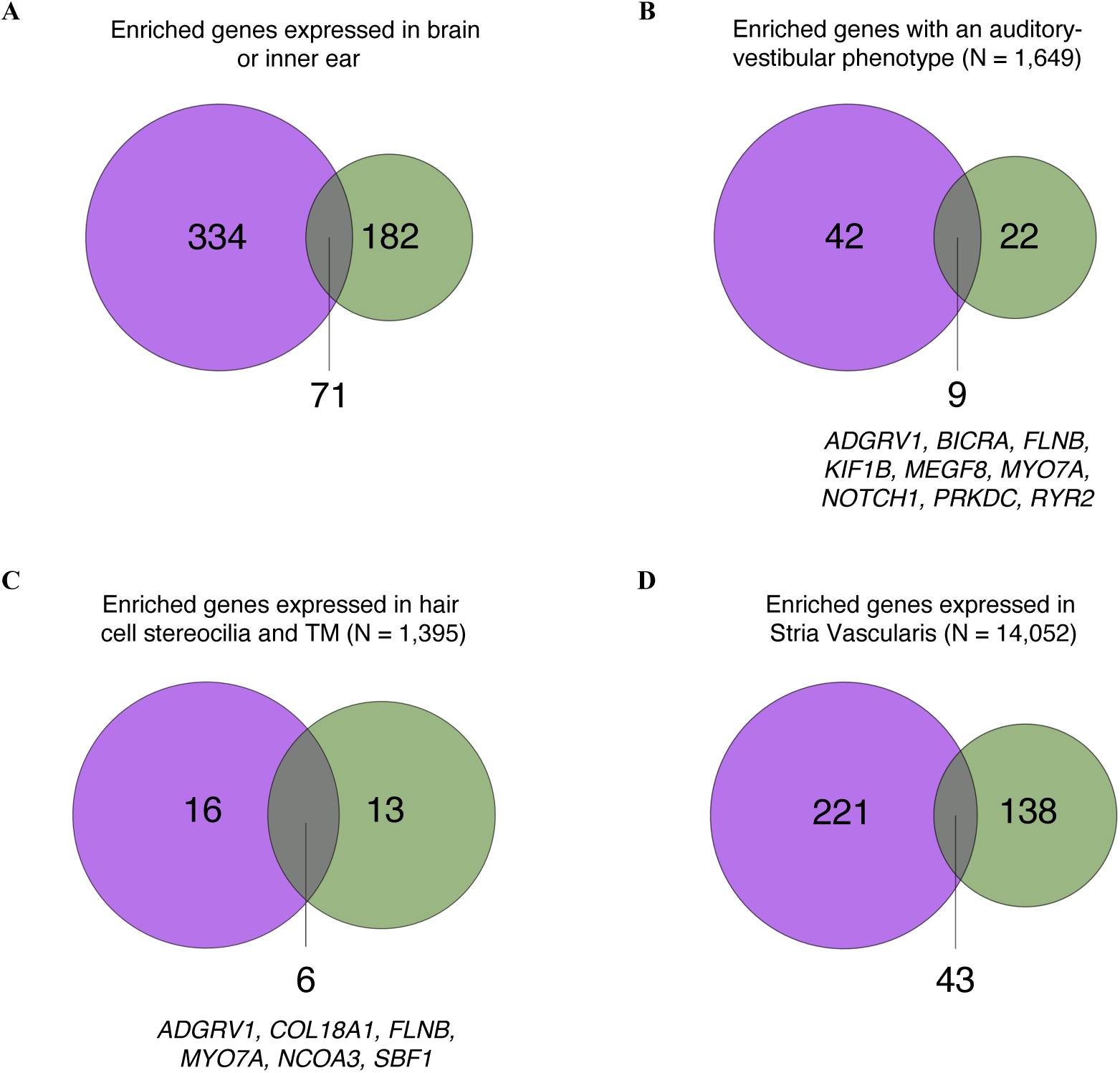
Venn diagrams of enriched genes obtained from the SMD (green) and FMD (purple) GBA. **A**: Using all enriched genes expressed in brain or inner ear. **B**: Using enriched genes expressed in brain or inner ear and with an associated auditory-vestibular phenotype. **C**: Using enriched genes that encode for proteins found in hair cell stereocilia and tectorial membrane (TM). **D**: Using enriched genes expressed in stria vascularis cells.

For the GBA with LoF variants, the enriched genes shared between SMD and FMD were *SYNGAP1*, *SKA3*, and *BTNL8*.

To understand the implications of SMD and FMD enriched genes, several enrichment functional analyses were performed. Regarding the functional analysis with the Cellular Component (CC) ontology, 13 significant GO terms (FDR < 0.05) were obtained for both SMD and FMD (Fig. 2e, Fig. 3e, Table S19 and Table S20). The overlapping terms were: “*Extracellular matrix*” (GO:0031012), “*Cytoskeleton*” (GO:0005856), “*Voltage-gated sodium channel complex*” (GO:0001518), and “*Kinesin complex*” in the microtubule (GO:0005871). Using the Molecular Function (MF) ontology of the GO database, 24 and 21 significant terms (p-value<0.05) were obtained for SMD and FMD, respectively (Fig. 2e, Fig. 3e, Table S21 and Table S22). Of note, the enriched terms include “*Microtubule motor activity*” (GO:0003777), “*Calcium ion binding*” (GO:0005509), and “*Actin filament binding*” (GO:0051015).

### Cellular distribution of MD enriched genes in the inner ear

Among the enriched genes in the SMD cohort, 19 genes are expressed in the hair cell whose resulting proteins are found in the stereocilia and TM (N = 1392, 1.36% of the total proteins) (Fig. 4c), and 181 genes are expressed in the stria vascularis (N = 14,052, 1.29% of the total expressed genes) (Fig. 4d). Similarly, for the FMD cohort, 22 genes are expressed in the hair cell bundles and TM (1.72% of the total proteins) (Fig. 4c), and 264 genes in the stria vascularis (1.89% of the total expressed genes) (Fig. 4d). In the case of enriched genes whose proteins are found in stereocilia, variants in these genes are found in 82.9% (238 out of 287) of SMD patients, and in 83.9% of FMD patients (78 out of 93). Concerning the enriched genes expressed in the stria vascularis, variants were found in all SMD and FMD patients.

Among them, 6 enriched genes were shared by both cohorts (*ADGRV1* (HGNC:17416), *COL18A1* (HGNC:2195), *FLNB* (HGNC:3755), *MYO7A* (HGNC:7606), *NCOA3* (HGNC:7670), and *SBF1* (HGNC:10542)) (Fig. 4c) in the case of those genes expressing proteins present in the hair cell stereocilia and TM. In addition, 43 enriched genes, shared in SMD and FMD were expressed in the stria vascularis cells (Table S23) (Fig. 4d).

## Discussion

This study compares the distribution of rare variants in SMD and FMD and supports an overload of rare missense and LoF variants that may contribute to MD development. The identification of different and overlapping genes between SMD and FMD confirms a complex genetic architecture and underlines the relevance of understanding both forms to develop more effective molecular diagnostic strategies.

In SMD cohort, a significant variant enrichment in 779 genes compared to global, NFE and Spanish reference populations was identified. After filtering, we found 31 genes, with rare variants in more than 9 patients, with phenotypic annotations related to the inner ear. Most of SMD patients (94%) had rare variants in these 31 genes, suggesting a higher predisposition to suffer MD. Notably, 19 of these genes are expressed in the hair cell stereocilia, highlighting their potential role in the pathophysiology of SMD. In addition, 181 enriched genes are found expressed in stria vascularis cells.

Among the genes enriched in SMD, *ADGRV1*, *PCDH15* and *MYO7A* stand out for their location at the stereocilia in the tip links and ankle links. Variants in these genes could contribute to a disorganization and lack of coordination in the stereocilia that would lead to abnormal gating of the MET channel, resulting in prolonged hair cell depolarization, vertigo and hearing impairment ^13^. Besides, *MYO7A* and *PCDH15* have been previously associated with FMD ^8^. Other noteworthy genes include *LOXHD1*, a gene described for DFNB77, which is located along the plasma membrane of hair cell stereocilia and is required for the mechanotransduction process ^14^; *CHRNA10*, which codes for an ionotropic receptor that modulates auditory stimuli and was found associated with hearing loss in the UK Biobank cohort ^15,16^; and *COL11A2*, associated with DFNA53 and DFNB13 ^17^. Besides, 16 genes (*ADGRV1, ARID1B, BICRA, CDK5RAP2, COL11A2, DMD, DMXL2, FLNB, LRP5, MEGF8, MYO7A, NIPBL, PCDH15, PRKDC, VCL, ZNF469*) have been previously associated with SNHL in HPO and 10 are located at cell-cell junctions according to CC gene ontology, supporting the formation of endolymphatic hydrops observed in some MD patients ^18,19^.

Similarly, in the FMD cohort, a significant enrichment of variants with a moderate impact was observed in 740 genes. After filtering and prioritizing, 51 enriched genes with inner ear phenotypic annotations were identified. Most of FMD patients (95.7%) had variants in these 51 genes, also suggesting a high relevance of these genes in the MD development. Among all enriched genes, 22 genes are expressed in hair cell stereocilia and TM, supporting their involvement in FMD under the hypothesis of fragile stereocilia could be destabilized and may provoke vertigo attacks and hearing loss ^9^. Besides, 264 genes are found expressed in stria vascularis cells.

Previously FMD-associated genes *MYO7A* and *TECTA* are included among the enriched genes ^8,10^. In addition, *ADGRV1, COL2A1, COL4A4, KARS1, LARS2, MYH14, OTOF, TNC,* and *TPRN*, have been described for several forms of dominant and recessive hearing loss according to OTOscope v9 and 19 additional genes have been associated with SNHL in HPO.

All FMD and SMD patients had variants with a moderate impact on the enriched genes, which could contribute to the different MD expressivity. This would suggest that SMD patients present a recessive type of inheritance. Furthermore, the presence of multiple variants in those genes in several patients suggests a polygenic inheritance that may contribute to disease variability in both FMD and SMD.

Subsequently, if the genetic architecture of SMD and FMD is compared, an overlap in 71 enriched genes could be observed. Common genes, such as *ADGRV1, BICRA, FLNB, KIF1B, MYO7A, NOTCH1,* and *PRKDC*, could indicate common genetic pathways involved in both forms of the disease and a possible hidden inheritance in SMD. These mechanisms could be related to both an aberrant hair cell stereocilia structure ^9^ and alterations in the stria vascularis in terms of cell-cell junction ^19^, for example. However, our results support additional pathogenic mechanisms in SMD and FMD, according to the number of variants and enriched genes found.

The concept of polygenic inheritance in SMD has been also supported in a large cohort of unilateral MD patients (N = 527), that reported 481 genes were involved in hearing, balance, and cochlear function, as well as cell-cell adhesion and extracellular matrix organization ^19^. Comparing with this study, 48 SMD (6 with an auditory-vestibular phenotype) and 56 FMD genes (11 with an auditory-vestibular phenotype) of the total number of enriched genes were replicated. Combining the genes enriched in SMD and FMD, a total of 86 genes were replicated (Table S24). In the functional analysis, pathways related to proteins present in the extracellular matrix and cell-cell junctions were also identified.

Our study has several limitations. First, we have used stringent filters to shorten the MD candidate gene list, excluding genes in the FLAGS gene list (N=100) ^20^. These genes have a large coding sequence, exhibit lower evolutionary pressure and have been more frequently associated with rare pathological phenotypes, reporting rare variants possibly with a functional impact ^20^. Two genes that could be relevant in the pathogenesis of MD and that show an enrichment of rare variants in both SMD and FMD cohorts were found in FLAGS list. These genes are *MYO15A* and *USH2A*. In the case of *MYO15A*, 42 variants have been found in 61 patients with SMD, and 21 variants in 19 patients with FMD. This gene is associated with DFNB3 and codes for myosin XVa, a protein necessary for the organization of actin in auditory and vestibular hair cells. It plays a role in their maturation and in the formation of stereocilia, through the contribution of Whirlin proteins to the upper part of these structures ^21^. Regarding *USH2A*, 45 variants have been identified in 64 patients with SMD, and 20 variants in 16 patients with FMD. This gene is implicated in Usher syndrome 2A and codes for Usherin, a protein required for the formation of ankle links. These links are formed together with other proteins, including Myosin VIIa, Adhesion G-protein coupled receptor V1, PDZ domain-containing protein 7, Vezatin, Whirlin, and Vlgr1. These proteins help connect the growing stereocilia in developing hair cells ^22^.

Second, this study focuses on exonic variants, which probably does not cover all the potential molecular mechanisms underlying the onset of MD, and hence, the use of whole genomes could help to carry out a deeper investigation of the MD genetic architecture and its sporadic and familial forms.

Third, GBA identifies statistical associations rather than causality. To reduce incidental findings, we focused on genes with known auditory and vestibular function; however, additional functional studies are needed to confirm their pathogenic role in MD.

Finally, it would be interesting to analyze variants in genes involved in the immune response, as they could be contributing to the MD development in those patients whose etiology seems to be autoimmune or autoinflammatory.

## Conclusion

The genetic structure of sporadic and familial MD according to the distribution of rare variants is different, with some overlapping genes, such as *ADGRV1* and *MYO7A.* However, SMD and FMD have an overload of missense variants in stria vascularis and hair cell stereocilia genes that suggests a multiallelic, recessive inheritance pattern. The contribution of LoF variants to either SMD or FMD is lower compared to missense variants, but it seems to be more relevant in stria vascularis genes in SMD.

## Materials and Methods

### Patient recruitment

The recruitment of Meniere Disease (MD) patients was based on the diagnostic criteria established by the International Classification Committee for Vestibular Disorders of the Barany Society ^1^. A total of 454 patients with MD were recruited in Spain. Among them, 167 were considered as FMD with one or more first-degree relatives affected by MD from 93 different unrelated families, and 287 as SMD.

The protocol of this study was approved by the local ethics committee (MS/2014/02, Institutional Review Board for Clinical Research, Universidad de Granada, Spain) and all the subjects signed a written informed consent to donate biological samples. The research was performed following the principles of the Declaration of Helsinki revised in 2013.

### Exome sequencing

Sample collection, DNA extraction and Exome Sequencing (WES) was performed following a previously published protocol ^8^.

### Bioinformatic analysis

Sarek Nextflow pipeline, included in NF-Core ^23^, was utilized to perform the exome reference alignment, mapping to the GRCh38/hg38 human reference genome, base quality score recalibration (BSQR), variant calling and quality filtering. Genetic variants were called using the Haplotypecaller tool from GATK ^24^. The VCF files were normalized using the *norm* function from BCFtools and filtered according to the criteria employed by gnomAD database ^25^: Allele balance (AB) ≥ 0.2 and AB ≤ 0.8 (for heterozygous genotypes only), genotype quality (GQ) ≥ 20, and depth (DP) ≥ 10 (5 for haploid genotypes on sex chromosomes). Afterwards, using the BCFtools *merge* function, a MD variant dataset was created. Subsequently, variant quality filtering was conducted using Variant Quality Score Recalibration (VQSR), calculating the VQSLOD score. Variants were filtered according to the VQSLOD value, using the first tranche (90), which corresponds to a 90% truth sensitivity threshold. This cutoff was selected to balance sensitivity and specificity, prioritizing the detection of rare variants while reducing false positives.

Variants were annotated using Ensembl Variant Effect Predictor (VEP) ^26^. Variants with high or moderate impact on the protein for FMD and SMD, according VEP classification, were kept separately for further analyses. Two independent databases were used to obtain the variant’ allelic frequencies (AF) in reference populations. AF for non-Finish European (NFE) population (N=32,299) and for the global population (N=71,702) were retrieved from gnomAD database v.3.0 ^25^; and AF for the Spanish population (N=2,071) from Collaborative Spanish Variant Server (CSVS) ^27^.

To highlight genes associated with SMD and FMD, a Gene Burden Analysis (GBA) was performed using the SMD and FMD cohort as previously described ^28^. Only one individual from each family was selected, whenever possible according to the lowest age of onset and/or from the last generation. Variants with an AF < 0.01 in the CSVS, gnomAD NFE, and gnomAD global databases were retained.

For each reference population, the GBA was performed independently. For each variant, we retrieve the allele count (AC) and allele number (AN) in both cases and controls. The number of wild-type alleles (WT) was computed as WT = AN − AC. To obtain a gene-level estimate, AC and WT were summed across all variants within each gene. The following statistics were calculated:

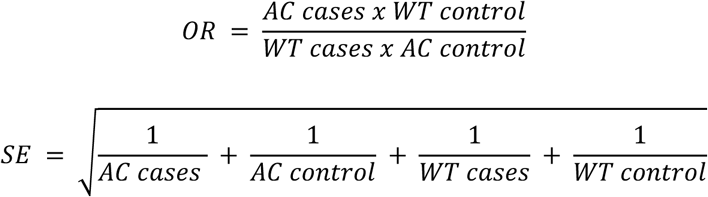

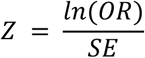

The two-tailed p-value (*p*) was calculated using the complementary cumulative distribution function (CCDF), with the formula:

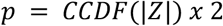

The p-value was corrected by false discovery rate (FDR), using the total number of genes:

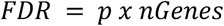

Finally, the 95 % CI was calculated as follows:

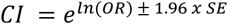

Variants do not present in the reference population were considered novel. In these cases, the AC was imputed as 0, and the AN was estimated as the average AN of all variants in the same gene.

Statistically significant genes (adjusted p-value < 0.05) and an OR ≥ 1 in the three comparisons with each reference population were considered enriched in MD.

### Gene prioritization

Genes with a high mutation rate (FLAGS) ^20^ were removed based on the top 100 genes listed. In addition, genes not expressed in the inner ear or brain were excluded. In order to obtain the genes expressed in the inner ear or in the brain, RNAseq data from several resources was used: 1) RNAseq data from inner ear (hair cells and non-hail cells) of P0 and adult mice from Expression Analysis Resource (gEAR) database ^29–31^, 2) RNAseq data from P30, M9 and M26 mice stria vascularis ^32^, 3) microarray RNA from P0 mouse Spiral Ganglion Neuron (SGN) and Vestibular Ganglion Neuron (VGN) from Shared Harvard Inner-Ear Laboratory Database (SHIELD) ^33^, 4) RNAseq data from three adult human cochleae, an ampulla, a saccule and an utricle from patients without hearing loss ^34^, and 5) RNAseq data from human brain tissues from Genotype-Tissue Expression (GTEx) project V8 ^35^. In addition, P3-P5 and P21-P25 mouse utricle proteomics data have been used to select those proteins present in hair cell stereocilia ^36^.

Genes with variants observed in at least 3% of individuals, even if each individual carried a different variant, were selected and ranked for further analysis. In addition, candidate genes in SMD and FMD were prioritized for their phenotypic association with hearing and balance through annotation with the Human Phenotype Ontology (HPO) ^37^ and Mouse Genomics Informatics (MGI) ^38^ databases. Genes annotated with any of the terms described in Table S25 was prioritized.

### Functional analysis

A functional enrichment analysis was carried out to discover the predominant localization and molecular functions related to the candidate genes. The enrichment analyses were performed via a hypergeometric test utilizing GeneCodis4 tool ^39^ and the Cellular Component (CC) and Molecular Function (MF) databases from Gene Ontology (GO) ^40^.

### Statistical analysis

The number of variants identified in each patient was compared between the FMD and SMD cohorts. The normality of the data distribution was assessed using the Shapiro-Wilk test. As the number of variants per patient did not follow a normal distribution, the non-parametric Mann-Whitney test was used. To account for multiple comparisons, the resulting p-values were adjusted using the Bonferroni correction method.

## Data Availability

The data that support the findings of this study are available from the corresponding author, (J.A.L-E.), upon request.

## Author Information

Conceptualization: J.A.L-E.; Investigation: A.M.P-P., A.G-M., A.E-B., P.R-B. & P.P-C.; Formal Analysis: A.M.P-P, A.E-B. & A.G-M.; Software: A.M.P-P. & A.E-B.; Supervision: J.A.L-E.; Writing-original draft: A.M.P-P. & J.A.L-E.; Writing-review and editing: A.M.P-P., A.G-M., A.E-B., P.R-B., P.P-C. & J.A.L-E. All authors approved the final manuscript.

## Conflict of Interest

The authors declare no conflicts of interest.

## Funding

Alberto M. Parra-Perez is supported by Andalusian University, Research and Innovation Department (Grant PREDOC2021/00343). Alvaro Gallego-Martinez has been funded by the Andalusian University, Research, and Innovation Department (Grant DOC_01677) and Andalusian Health Department (Grant PI-0266-2021 GEN4PHEN), Cures Within Reach and the Knight family. Jose A Lopez-Escamez has received funds to support research on genetics in MD from The University of Sydney (Grant K7013_B341), Andalusian Health Department (Grant PI027-2020), Asociación Sindrome de Meniere España (ASMES) and Meniere’s Society, UK. Patricia Perez-Carpena has received funds from Instituto de Salud Carlos III (Grant PI22/01838).

## Ethics Declaration

This study has been approved by a local ethics committee (MS/2014/02, Institutional Review Board for Clinical Research, Universidad de Granada, Spain). Written informed consent for publication was obtained from all patients. Data have been deidentified for publication. Studies were performed in accordance with the Declaration of Helsinki.

## Acknowledgments

Alberto M. Parra-Perez is enrolled in the Biomedicine Ph.D. program at Universidad de Granada, Spain, and this work is part of his doctoral thesis. The authors thank the patients with MD for their enthusiastic participation, and clinicians for their contribution to patient identification and clinical characterization. In addition, we would like to thank all former members of the Otology and Otoneurology CTS495 group for years of research and contributing to the sample processing and the development of the bioinformatics methods used in this work.

## Notes

### Competing Interest Statement

The authors have declared no competing interest.

### Author Declarations

Institutional Review Board for Clinical Research of Universidad de Granada, Spain, gave ethical approval for this work (MS/2014/02).

## References

1. Lopez-Escamez JA, Carey J, Chung WH, et al. Diagnostic criteria for Ménière’s disease. Journal of Vestibular Research. 2015;25(1):1–7. doi:10.3233/VES-150549

2. Ohmen JD, White CH, Li X, et al. Genetic Evidence for an Ethnic Diversity in the Susceptibility to Ménière’s Disease. Otology & Neurotology. 2013;34(7):1336. doi:10.1097/MAO.0b013e3182868818

3. Frejo L, Soto-Varela A, Santos-Perez S, et al. Clinical Subgroups in Bilateral Meniere Disease. Front Neurol. 2016;7:182. doi:10.3389/fneur.2016.00182

4. Flook M, Rojano E, Gallego-Martinez A, et al. Cytokine profiling and transcriptomics in mononuclear cells define immune variants in Meniere Disease. Genes Immun. Published online February 23, 2024. doi:10.1038/s41435-024-00260-z

5. Requena T, Espinosa-Sanchez JM, Cabrera S, et al. Familial clustering and genetic heterogeneity in Meniere’s disease. Clin Genet. 2014;85(3):245–252. doi:10.1111/cge.12150

6. Morrison AW. Anticipation in Menière’s disease. The Journal of laryngology and otology. 1995;109(6):499–502.

7. Roman-Naranjo P, Gallego-Martinez A, Soto-Varela A, et al. Burden of Rare Variants in the OTOG Gene in Familial Meniere’s Disease. Ear and hearing. 2020;41(6):1598–1605. doi:10.1097/AUD.0000000000000878

8. Roman-Naranjo P, Moleon MDC, Aran I, et al. Rare coding variants involving MYO7A and other genes encoding stereocilia link proteins in familial meniere disease. Hear Res. 2021;409:108329. doi:10.1016/j.heares.2021.108329

9. Parra-Perez AM, Lopez-Escamez JA. Types of Inheritance and Genes Associated with Familial Meniere Disease. JARO. Published online April 6, 2023. doi:10.1007/s10162-023-00896-0

10. Roman-Naranjo P, Parra-Perez AM, Escalera-Balsera A, et al. Defective α-tectorin may involve tectorial membrane in familial Meniere disease. Clin Transl Med. 2022;12(6):e829. doi:10.1002/ctm2.829

11. Gallego-Martinez A, Requena T, Roman-Naranjo P, Lopez-Escamez JA. Excess of Rare Missense Variants in Hearing Loss Genes in Sporadic Meniere Disease. Front Genet. 2019;10:76. doi:10.3389/fgene.2019.00076

12. Gallego-Martinez A, Requena T, Roman-Naranjo P, May P, Lopez-Escamez JA. Enrichment of damaging missense variants in genes related with axonal guidance signalling in sporadic Meniere’s disease. Journal of medical genetics. 2020;57(2):82–88. doi:10.1136/jmedgenet-2019-106159

13. Senofsky N, Faber J, Bozovic D. Vestibular Drop Attacks and Meniere’s Disease as Results of Otolithic Membrane Damage—A Numerical Model. J Assoc Res Otolaryngol. 2023;24(1):107–115. doi:10.1007/s10162-022-00880-0

14. Trouillet A, Miller KK, George SS, et al. Loxhd1 Mutations Cause Mechanotransduction Defects in Cochlear Hair Cells. J Neurosci. 2021;41(15):3331–3343. doi:10.1523/JNEUROSCI.0975-20.2021

15. Elgoyhen AB, Katz E, Fuchs PA. The Nicotinic Receptor of Cochlear Hair Cells: A Possible Pharmacotherapeutic Target? Biochem Pharmacol. 2009;78(7):712–719. doi:10.1016/j.bcp.2009.05.023

16. Lewis MA, Schulte BA, Dubno JR, Steel KP. Investigating the characteristics of genes and variants associated with self-reported hearing difficulty in older adults in the UK Biobank. BMC Biol. 2022;20:150. doi:10.1186/s12915-022-01349-5

17. Chakchouk I, Grati M, Bademci G, et al. Novel mutations confirm that COL11A2 is responsible for autosomal recessive non-syndromic hearing loss DFNB53. Mol Genet Genomics. 2015;290(4):1327–1334. doi:10.1007/s00438-015-0995-9

18. van der Lubbe MFJA, Vaidyanathan A, Van Rompaey V, et al. The “hype” of hydrops in classifying vestibular disorders: a narrative review. J Neurol. 2020;267(1):197–211. doi:10.1007/s00415-020-10278-8

19. Fisch KM, Rosenthal SB, Mark A, et al. The genomic landscape of Ménière’s disease: a path to endolymphatic hydrops. BMC Genomics. 2024;25(1):646. doi:10.1186/s12864-024-10552-3

20. Shyr C, Tarailo-Graovac M, Gottlieb M, Lee JJY, Karnebeek C van, Wasserman WW. FLAGS, frequently mutated genes in public exomes. Published online December 2014:1–14.

21. Rehman AU, Bird JE, Faridi R, et al. Mutational Spectrum of MYO15A and the Molecular Mechanisms of DFNB3 Human Deafness. Hum Mutat. 2016;37(10):991–1003. doi:10.1002/humu.23042

22. Michalski N, Michel V, Bahloul A, et al. Molecular Characterization of the Ankle-Link Complex in Cochlear Hair Cells and Its Role in the Hair Bundle Functioning. J Neurosci. 2007;27(24):6478–6488. doi:10.1523/JNEUROSCI.0342-07.2007

23. Garcia M, Juhos S, Larsson M, et al. Sarek: A portable workflow for whole-genome sequencing analysis of germline and somatic variants. F1000Res. 2020;9:63. doi:10.12688/f1000research.16665.2

24. O’Connor BD, van der Auwera G. Genomics in the Cloud: Using Docker, GATK, and WDL in Terra. O’Reilly Media, Incorporated; 2020. https://books.google.es/books?id=wwiCswEACAAJ

25. Chen S, Francioli LC, Goodrich JK, et al. A genome-wide mutational constraint map quantified from variation in 76,156 human genomes. Published online October 10, 2022:2022.03.20.485034. doi:10.1101/2022.03.20.485034

26. McLaren W, Gil L, Hunt SE, et al. The Ensembl Variant Effect Predictor. Genome Biol. 2016;17:122. doi:10.1186/s13059-016-0974-4

27. Peña-Chilet M, Roldán G, Perez-Florido J, et al. CSVS, a crowdsourcing database of the Spanish population genetic variability. Nucleic Acids Research. 2021;49(D1):D1130–D1137. doi:10.1093/nar/gkaa794

28. Walsh R, Mazzarotto F, Whiffin N, et al. Quantitative approaches to variant classification increase the yield and precision of genetic testing in Mendelian diseases: the case of hypertrophic cardiomyopathy. Genome Medicine. 2019;11(1):5. doi:10.1186/s13073-019-0616-z

29. Elkon R, Milon B, Morrison L, et al. RFX transcription factors are essential for hearing in mice. Nat Commun. 2015;6(1):8549. doi:10.1038/ncomms9549

30. Li Y, Liu H, Giffen KP, Chen L, Beisel KW, He DZZ. Transcriptomes of cochlear inner and outer hair cells from adult mice. Sci Data. 2018;5(1):180199. doi:10.1038/sdata.2018.199

31. Liu H, Chen L, Giffen KP, et al. Cell-Specific Transcriptome Analysis Shows That Adult Pillar and Deiters’ Cells Express Genes Encoding Machinery for Specializations of Cochlear Hair Cells. Front Mol Neurosci. 2018;11. doi:10.3389/fnmol.2018.00356

32. Liu H, Giffen KP, Chen L, et al. Molecular and cytological profiling of biological aging of mouse cochlear inner and outer hair cells. Cell Rep. 2022;39(2):110665. doi:10.1016/j.celrep.2022.110665

33. Shen J, Scheffer DI, Kwan KY, Corey DP. SHIELD: an integrative gene expression database for inner ear research. Database (Oxford). 2015;2015:bav071. doi:10.1093/database/bav071

34. Schrauwen I, Hasin-Brumshtein Y, Corneveaux JJ, et al. A comprehensive catalogue of the coding and non-coding transcripts of the human inner ear. Hear Res. 2016;333:266–274. doi:10.1016/j.heares.2015.08.013

35. Lonsdale J, Thomas J, Salvatore M, et al. The Genotype-Tissue Expression (GTEx) project. Nat Genet. 2013;45(6):580–585. doi:10.1038/ng.2653

36. Krey JF, Sherman NE, Jeffery ED, Choi D, Barr-Gillespie PG. The proteome of mouse vestibular hair bundles over development. Sci Data. 2015;2:150047. doi:10.1038/sdata.2015.47

37. Gargano MA, Matentzoglu N, Coleman B, et al. The Human Phenotype Ontology in 2024: phenotypes around the world. Nucleic Acids Research. 2024;52(D1):D1333–D1346. doi:10.1093/nar/gkad1005

38. Baldarelli RM, Smith CL, Ringwald M, Richardson JE, Bult CJ, Mouse Genome Informatics Group. Mouse Genome Informatics: an integrated knowledgebase system for the laboratory mouse. Genetics. 2024;227(1):iyae031. doi:10.1093/genetics/iyae031

39. Garcia-Moreno A, López-Domínguez R, Villatoro-García JA, et al. Functional Enrichment Analysis of Regulatory Elements. Biomedicines. 2022;10(3):590. doi:10.3390/biomedicines10030590

40. The Gene Ontology Consortium, Aleksander SA, Balhoff J, et al. The Gene Ontology knowledgebase in 2023. Genetics. 2023;224(1):iyad031. doi:10.1093/genetics/iyad031

